# Declines and pronounced state disparities in prescription opioid distribution in the United States

**DOI:** 10.1101/2021.12.02.21266660

**Authors:** Joshua D. Madera, Amanda E. Ruffino, Adriana Feliz, Kenneth L. McCall, Corey S. Davis, Brian J. Piper

**Affiliations:** Geisinger Commonwealth School of Medicine, Scranton, PA; University of New England, Portland, ME; Network for Public Health Law, Los Angeles, CA; Center for Pharmacy Innovation and Outcomes, Forty Fort, PA

## Abstract

**Background:** The United States (US) opioid epidemic is a persistent and pervasive public health emergency that now claims the lives of over 100,000 Americans per year. There have been sustained efforts to reverse this crisis over the past decade, including a number of measures designed to decrease the use of prescription opioids for the treatment of pain. This study analyzed the changes in federal production quotas for prescription opioids and the distribution of prescription opioids for pain, and identified state-level differences between 2010 and 2019.

**Methods and Findings:** Data on opioid production quotas and distribution of ten prescription opioids (codeine, fentanyl, hydrocodone, hydromorphone, meperidine, methadone, morphine, oxycodone, oxymorphone, and tapentadol) for 2010 to 2019 were obtained from the Drug Enforcement Administration. Total opioid production quotas increased substantially from 2010 to 2013 before decreasing 41.5% from 2013 (87.6 morphine mg equivalent metric tons) to 2019 (51.3). The peak year for distribution of all ten prescription opioids was between 2010 and 2013, except for codeine (2015). The largest quantities of opioid distribution were observed in Tennessee (520.70 morphine mg equivalent or MME per person) and Delaware (251.45) in 2011 and 2019. There was a 52.0% overall decrease in opioid distribution per capita from 2010 to 2019, with the largest decrease in Florida (−61.6%) and smallest in Texas (−18.6%). The highest to lowest state ratio of total opioid distribution, corrected for population, decreased from 5.25 in 2011 to 2.78 in 2019. The mean 95^th^/5^th^ ratio was relatively consistent in 2011 (4.78 ±0.70) compared to 2019 (5.64±0.98). Southern states had the highest per capita distribution for eight of the ten opioids in 2019.

**Conclusions:** This study found a sustained decline in distribution of ten prescription opioids during the last half-decade. Distribution was non-homogeneous at the state level. Analysis of state-level differences revealed a three-fold difference in the 95^th^:5^th^ percentile ratio between states which was unchanged over the past decade. Production quotas did not correspond with the distribution, particularly in the 2010-2016 period. Future research focused on identifying factors contributing to the observed regional variability in opioid distribution could prove valuable to understanding, and potentially remediating, the pronounced disparities in prescription opioid-related harm in the US.

## Background

Drug overdoses, largely driven by opioids, recently exceeded one-hundred thousand in one-year, the highest number ever recorded [1]. Despite a pronounced decline in prescription opioid consumption over the past decade, prescription opioids continue to have a strong presence throughout the US. The opioid prescription per capita increased by 7.3% between 2007 and 2012 with larger elevations in pain medicine (8.5%) and physical medicine/rehabilitation (12.0%) [2].

This profile has since reversed. Following an overall peak in 2011, there has been a gradual decline in opioid distribution in the US [3,4]. Despite these reductions, the amount of opioids prescribed remains elevated, and in 2015 [5] was almost three-fold higher than 1999 [6]. The morphine milligram equivalent (MME) per capita in 2016 in the US was 1,124 [3], which is appreciably higher than other developed countries [7] or the US Territories [8]. Inter-state variability in opioid distribution rates was also observed in 2016. There was a five-fold difference between the lowest (North Dakota = 485) and highest (Rhode Island = 2,624) states [3]. Examination of individual opioids [9, 10] revealed a three-fold state-level difference for fentanyl [11], eighteen-fold for meperidine [12], and twenty-fold for buprenorphine [13].

This study aimed to identify more up to date and conclusive patterns in the production quotas for and distribution of ten prescription opioids used primarily for pain between 2010 and 2019. This range was chosen as it includes the implementation of legislative changes in most states intended to limit the distribution of opioids [11, 14] and is prior to the COVID-19 pandemic and associated drug shortages [15]. Correlations among state-level distribution of individual opioids were also determined. Identifying the states with the highest and lowest declines in opioid distribution as well as quantifying this variation may be useful to inform research into the correlates and causes of opioid-related harm as well as efforts to determine the effectiveness of efforts to reduce the impact of that harm.

## Method

### Procedures

Opioid production quota data were obtained from the US Drug Enforcement Administration’s (DEA) Production Quotas System [16, Supplemental Table 1], and opioid distribution data from their Automation of Reports and Consolidated Orders System (ARCOS). ARCOS is a publicly available reporting system that reports detailed and comprehensive drug information from manufacturing to distribution [17]. ARCOS data corresponds highly with dispensing of opioids as reported to state prescription drug monitoring programs [3, 18] and has been used in many prior investigations [3, 4, 8-15, 18, 19]. Information on ten Schedule II opioid prescription drugs used for the treatment of pain: codeine, fentanyl base, hydrocodone, hydromorphone, meperidine, methadone, morphine, oxycodone, oxymorphone, and tapentadol, were collected. Buprenorphine [13] was excluded because it is primarily employed for treatment of opioid use disorder (OUD). All procedures were approved by Geisinger and the University of New England’s Institutional Review Board.

### Data Analysis

The oral MME for each opioid drug was determined using the appropriate multipliers: codeine 0.15, fentanyl base 75, hydrocodone 1, hydromorphone 4, meperidine 0.1, methadone 8, morphine 1, oxycodone 1.5, oxymorphone 3, and tapentadol 0.4 [3, 4] and summed for all ten opioids across all 50 states. Methadone [9], when distributed for the treatment of OUD, was excluded. The weight in grams of each opioid was obtained from ARCOS from 2010 to 2019 to isolate the peak opioid distribution year. Heat maps demonstrating the percent change across all 50 states per capita were produced using JMP. The per capita distribution of each opioid drug for each state was calculated using the respective population estimate from the American Community Survey [20]. Ratios for the highest to lowest states and for the 95^th^ : 5^th^ and 70^th^ : 30^th^ percentiles were calculated using these values [21]. To calculate these ratios, the per capita distribution of each opioid drug was organized from largest to smallest value, and the 95th and 5th percentile values were found. Using these values, the aforementioned ratios were subsequently calculated. Spearman correlations were calculated between each opioid for per capita distribution. The standard error of the mean (SEM) depicted variability. States outside of a 95% confidence interval were identified. A Fisher r to Z transformation was completed to determine if the correlations differed between 2011 and 2019.

## Results

### Production

Examination of total opioid production quota data from 2010 (54.9 MME metric tons) to 2019 revealed a gradual increase from 2011 to 2012, followed by a steep increase from 2012 to 2013 (87.6 tons), which remained relatively steady until a pronounced decline from 2016 (85.2 tons) to 2017 (57.6 tons). There was a 39.8% reduction from 2016 to 2019 (51.3 tons). Meperidine (−60.0%), oxymorphone (−54.3%), morphine (−49.6%), fentanyl (−48.5%) and hydrocodone (−48.1%), exhibited pronounced declines. Oxycodone (−38.5%), codeine (−37.5%), hydromorphone (−32.9%), methadone (−30.1%), and tapentadol (−27.5%) had sizable but more modest reductions in opioid production quotas (Figure 1A).

**Figure 1.**
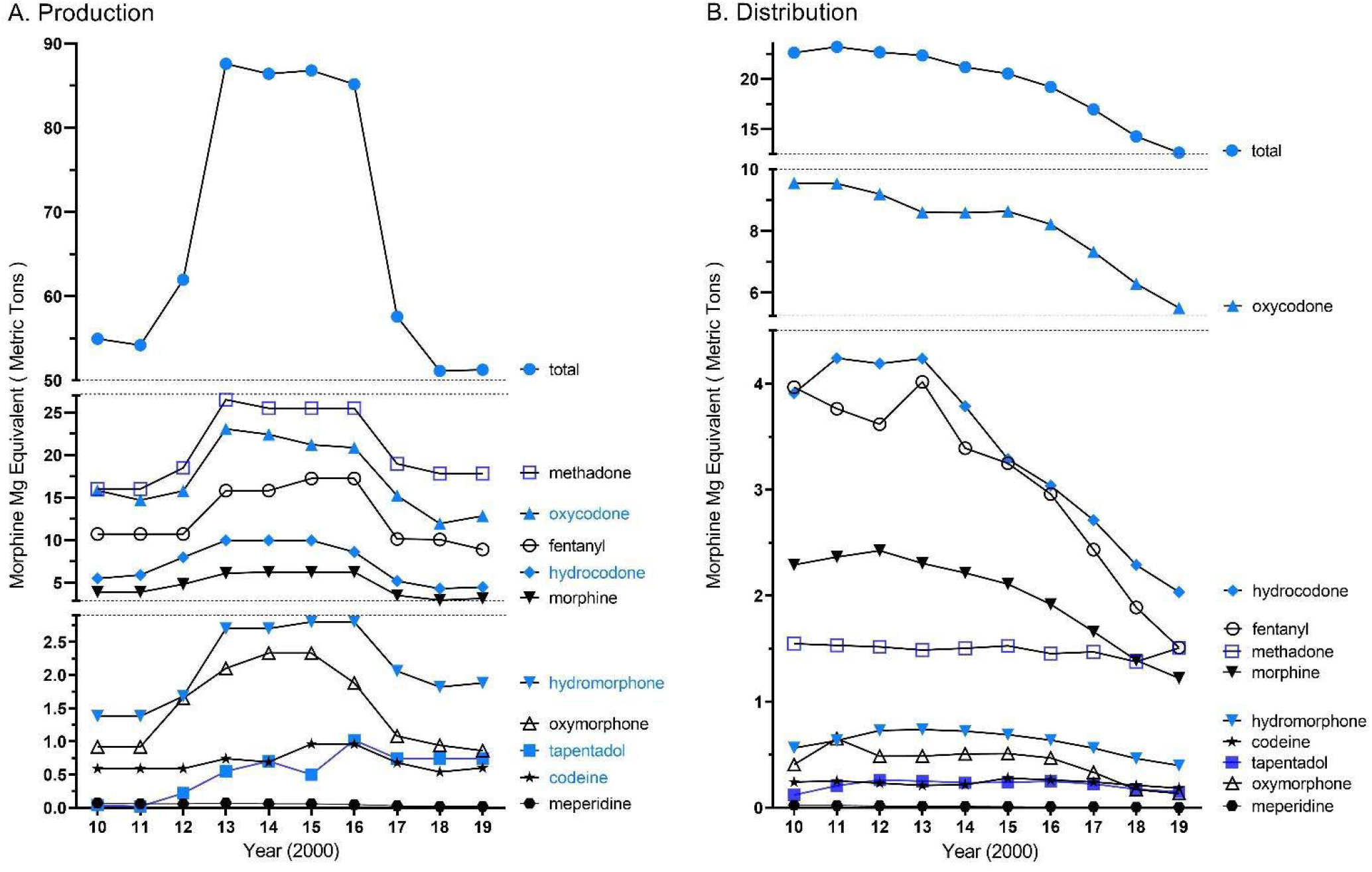
Total opioid production quotas (A) and distribution (B) in morphine mg equivalents (MME) as reported to the Drug Enforcement Administration across the United States from 2010 to 2019 of ten Schedule II opioids used primarily for pain.

### Distribution

Analysis of prescription opioid distribution from 2010 to 2019 identified 2011 as the peak year overall (Figure 1B). The peak for each drug was 2010 for methadone, oxycodone and meperidine; 2011 for hydrocodone and oxymorphone; 2012 for morphine and tapentadol; 2013 for hydromorphone and fentanyl; and 2015 for codeine. Since 2011 (23.2 MME metric tons), total opioid distribution for the ten opioids for pain has declined. From 2011 to 2019 (12.6 tons), a total decline in the opioid distribution of 45.4% was observed, with 2018 to 2019 showing an 11.3% decline.

Comparison of total opioid distribution per capita in 2011 (Supplemental Figure 1A) and 2019 (Supplemental Figure 1B) revealed an overall decrease (−51.96%), but also considerable state-level differences. In 2011, four states, Tennessee (520.70), Nevada (491.38), Delaware (476.15), and Florida (452.12), had significantly elevated levels of total opioid distribution relative to the national mean (284.34, *p* < 0.05, Figure 2A). However, only two-states, Alabama (251.45) and Delaware (238.71) were significantly above the national average (162.09) in 2019. Forty-three states had an MME per capita > 200 in 2011 relative to only six in 2019 (χ^2^(1) = 54.78, *p* < .0001). Four of these six were in the southeastern (KT, TN) or south-central (AL, OK) US. Analysis of individual opioid distribution trends revealed codeine to have the largest per capita increase (+143.09%) and oxycodone the largest decrease (−50.09%) from 2011 to 2019. Furthermore, there was a 3.31 fold difference between the highest (Florida = -61.61%) and lowest (Texas = -18.64%) states for the decline in opioid distribution. Figure 2B demonstrates the observed percent difference for total opioid distribution during this period and showed that other states with > 50% reductions were Nevada (−59.7%), Ohio (−57.4%), Tennessee (−55.2%), Maine (−54.4%), Oregon (−54.3%), and West Virginia (−50.9%).

**Figure 2.**
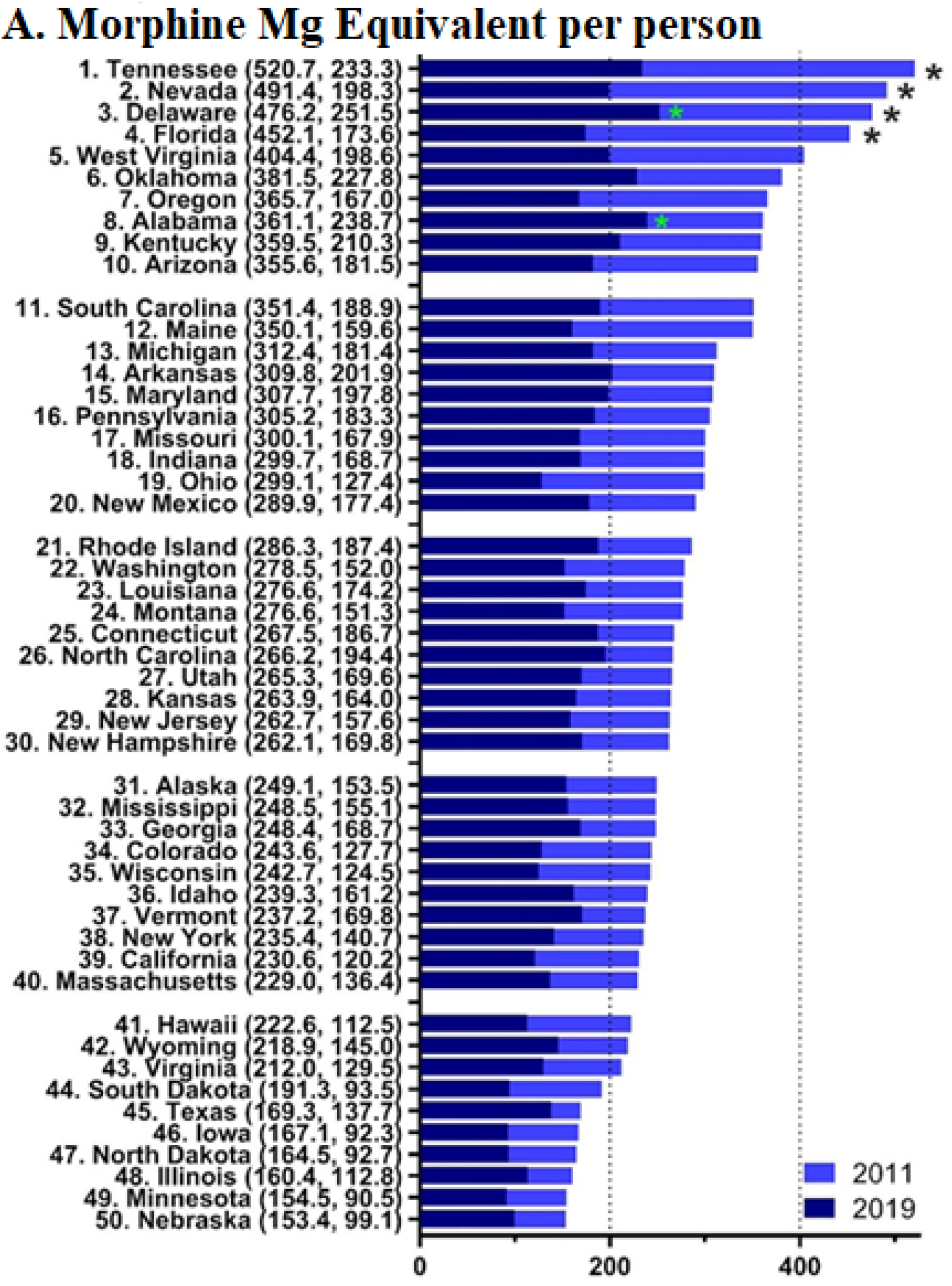
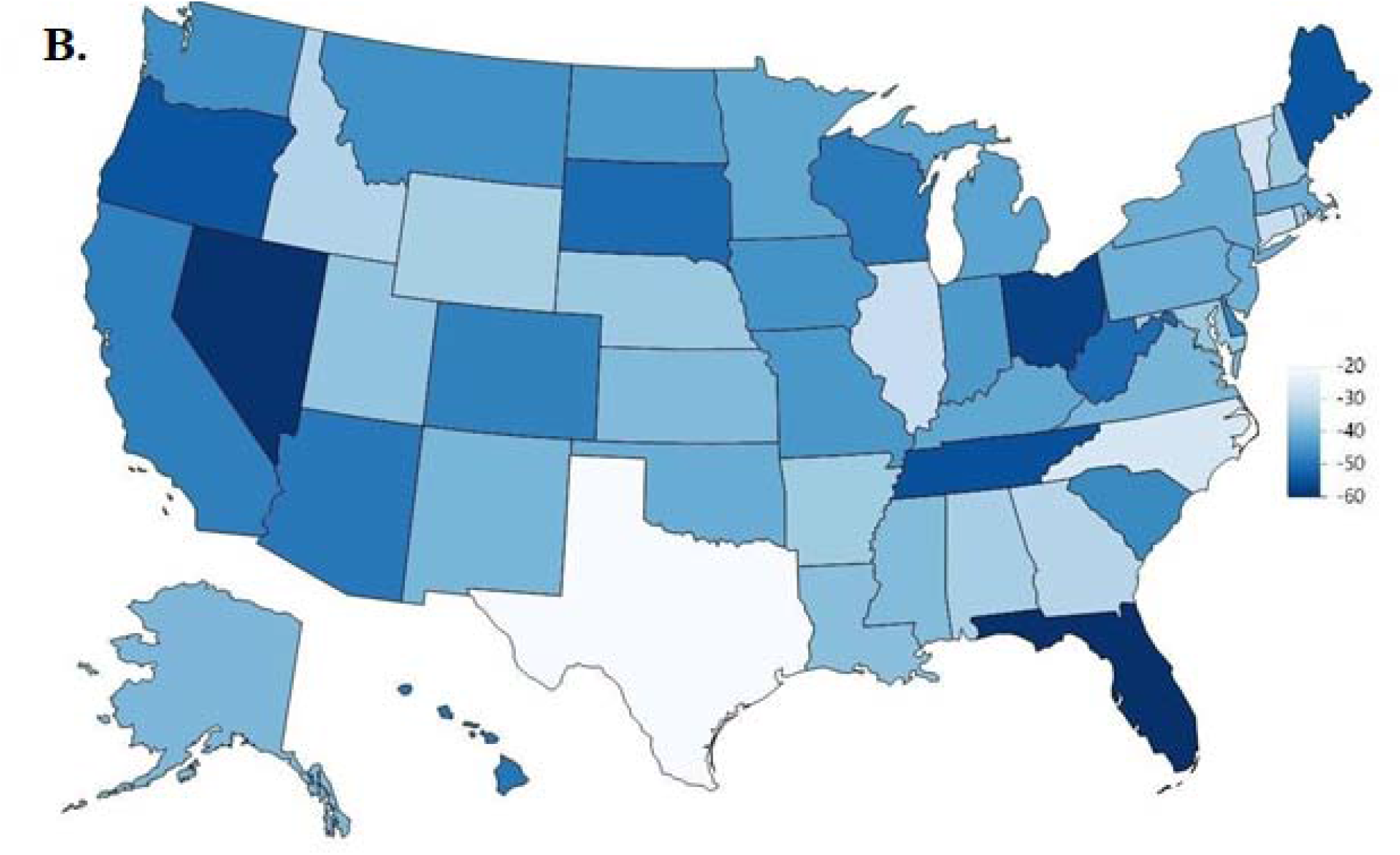
Per person distribution was organized from largest to smallest values for 2011 with the 2019 per person distribution superimposed (A) with the values in 2019 and 2011 in parentheses after each state (* *p* < .05 versus the average states in 2011 (black font) and 2019 (green font)). Heat map percent reduction in total opioid distribution from 2011 to 2019 (B).

Figure 3 shows that in 2011, the ratio of per capita MME distribution of opioids from the maximum to minimum states was highest for oxymorphone (27.4) followed by meperidine (19.6), tapentadol (14.0), oxycodone (13.6), methadone (11.2), hydrocodone (9.2), hydromorphone (4.6), morphine (4.2), codeine (4.0) and fentanyl (2.5, mean = 11.0 ±2.4). The ratio of per capita MME distribution of opioids from the maximum to minimum states for 2019 was again highest for oxymorphone (38.9), but instead was followed by methadone (30.3), meperidine (22.2), hydrocodone (13.4), tapentadol (12.3), codeine (9.8), oxycodone (6.1), hydromorphone (4.9), morphine (3.2), and again lowest for fentanyl (3.1) in 2019 (Figure 3B and D, mean = 14.4±3.7, *t*(9) = 1.45, *p* = .18). Eight states in the south had the highest per capita distribution for the ten opioids evaluated including Mississippi (fentanyl and hydromorphone), Tennessee (morphine and oxymorphone), Arkansas (meperidine), Alabama (hydrocodone), Delaware (oxycodone), and North Carolina (tapentadol) in 2019. The states with the lowest per capita distribution included Texas (oxycodone and morphine), Hawaii (fentanyl and hydromorphone), and Minnesota (oxymorphone and tapentadol) in 2019 (Supplemental Table 1).

**Figure 3.**
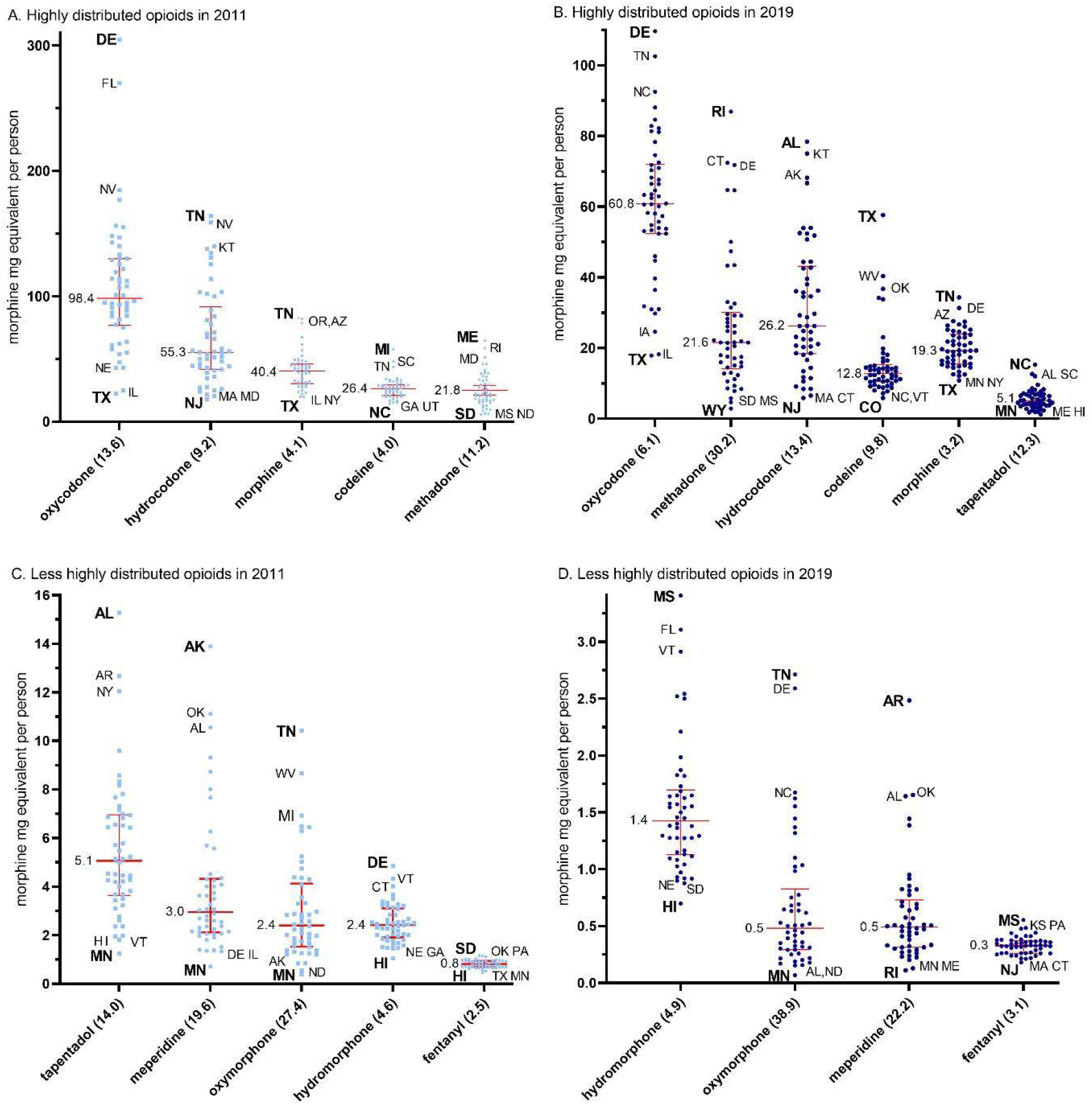
Median (+/- interquartile range) per person distribution for highly (A, B) and less highly (C, D) distributed prescription opioids by morphine mg equivalents per state in 2011 (A, C) and 2019 (B, D). The maximum to minimum ratio is listed in parentheses after each opioid. The median is written next to each (red) solid horizontal line. Abbreviations are shown for states outside of the 95% confidence interval with the highest and lowest states in **bold**.

Figure 4 shows the ratios between the 95^th^ : 5^th^ percentiles for the population corrected distribution of each Schedule II opioid for 2011 and 2019. The 95^th^ : 5^th^ percentile ratio identifies the degree of variation in distribution that each opioid experienced within a particular year across the US, where a larger ratio signifies a greater extent of variation. The 2011 95^th^ : 5^th^ percentile ratio was highest for oxymorphone (8.0) and meperidine (7.3), and smallest for codeine (2.4) and fentanyl (1.8). The 2019 95^th^ : 5^th^ percentile ratio ratio was highest for methadone (10.2) and oxymorphone (10.1) and lowest for morphine (2.21) and fentanyl (2.11) in 2019. A comparison of the 95^th^:5^th^ percentile ratios between 2011 (4.78 ±0.70) and 2019 (5.64 ±0.98) revealed a non-significant difference.

**Figure 4.**
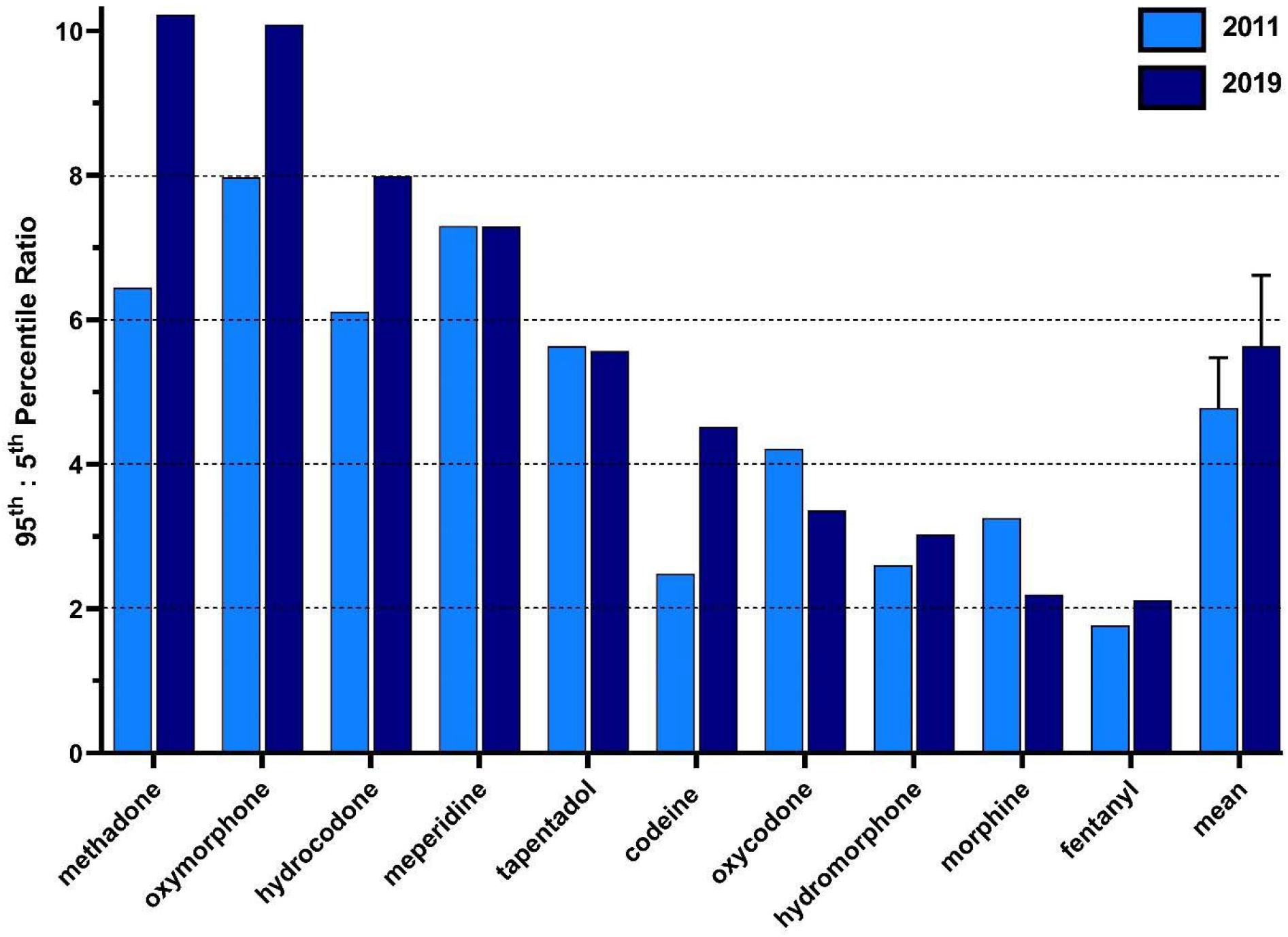
The 95^th^ to 5^th^ percentile ratio of the per person distribution per state for each prescription opioids in 2011 and 2019.

Table 1 shows correlations in the MME per capita per state between each opioid. Oxycodone had the most significant correlations with the other opioids in both 2011 and 2019. In 2011, there were three, mostly moderate (r = 0.35-0.69), correlations between oxycodone with morphine, oxymorphone, and methadone. Moreover, there were also moderate associations between hydrocodone and meperidine, and between oxymorphone and tapentadol. In 2019, there were three moderate correlations between oxycodone with morphine, oxymorphone, and methadone. There were also moderate correlations between hydrocodone with meperidine and codeine. Fischer r to Z statistics were calculated to identify differences in the strength of the correlations between opioids in 2011 relative to 2019. These analyses revealed a significant difference, i.e. an increase, in the magnitude of association between codeine and hydrocodone (*p* < 0.05), codeine and hydromorphone (*p* < 0.01), hydrocodone and methadone (*p* < 0.05), and methadone and oxycodone (p < 0.05). Comparisons between the averages of the correlations were then completed for each opioid with each other opioid. A significant difference was identified in the mean correlation between 2011 and 2019 for methadone (*p* < 0.05).

**Table 1.** Correlations for per capita state opioid distribution in 2011 and 2019. Boxes in blue and orange represent significant correlations between opioids of < 0.05 and < 0.01, respectively. * Significance between Z scores when comparing 2011 and 2019 correlations. ** Significance between mean correlations between 2011 and 2019.

| 2011 | A | B | C | D | E | F | G | H | I | Mean | Standard Error |
| --- | --- | --- | --- | --- | --- | --- | --- | --- | --- | --- | --- |
| codeine (A) | 1.00 |  |  |  |  |  |  |  |  | 0.15 | 0.05 |
| fentanyl (B) | 0.31 | 1.00 |  |  |  |  |  |  |  | 0.23 | 0.04 |
| hydrocodone (C) | 0.17* | 0.16 | 1.00 |  |  |  |  |  |  | 0.13 | 0.09 |
| hydromorphone (D) | 0.16* | 0.28 | -0.30 | 1.00 |  |  |  |  |  | 0.13 | 0.09 |
| meperidine (E) | -0.10 | 0.10 | 0.62 | -0.22 | 1.00 |  |  |  |  | 0.10 | 0.09 |
| methadone (F) | 0.08 | 0.06 | -0.15* | 0.40 | -0.20 | 1.00 |  |  |  | 0.16 | 0.10 |
| morphine (G) | 0.33 | 0.39 | 0.24 | 0.29 | 0.18 | 0.35 | 1.00 |  |  | 0.26 | 0.07 |
| oxycodone (H) | 0.19 | 0.26 | -0.12 | 0.44 | 0.02 | 0.66* | 0.50 | 1.00 |  | 0.29 | 0.09 |
| oxymorphone (I) | 0.25 | 0.36 | 0.32 | 0.18 | 0.16 | 0.35 | 0.28 | 0.50 | 1.00 | 0.32 | 0.04 |
| tapentadol (J) | 0.01 | 0.14 | 0.24 | -0.04 | 0.38 | -0.08 | -0.21 | 0.13 | 0.51 | 0.12 | 0.08 |

| 2019 | A | B | C | D | E | F | G | H | I | Mean | Standard Error |
| --- | --- | --- | --- | --- | --- | --- | --- | --- | --- | --- | --- |
| codeine (A) | 1.00 |  |  |  |  |  |  |  |  | 0.08 | 0.08 |
| fentanyl (B) | 0.07 | 1.00 |  |  |  |  |  |  |  | 0.15 | 0.06 |
| hydrocodone (C) | 0.54* | 0.33 | 1.00 |  |  |  |  |  |  | 0.12 | 0.12 |
| hydromorphone (D) | -0.31* | 0.14 | -0.33 | 1.00 |  |  |  |  |  | 0.06 | 0.08 |
| meperidine (E) | 0.17 | 0.29 | 0.58 | -0.07 | 1.00 |  |  |  |  | 0.17 | 0.09 |
| methadone (F) | -0.10 | -0.26 | -0.45* | 0.31 | -0.39 | 1.00 |  |  |  | -0.02** | 0.10 |
| morphine (G) | 0.13 | 0.31 | 0.26 | 0.30 | 0.26 | 0.13 | 1.00 |  |  | 0.25 | 0.05 |
| oxycodone (H) | 0.10 | 0.05 | 0.02 | 0.25 | 0.21 | 0.39* | 0.51 | 1.00 |  | 0.29 | 0.07 |
| oxymorphone (I) | 0.12 | 0.24 | 0.14 | 0.16 | 0.10 | 0.23 | 0.29 | 0.61 | 1.00 | 0.27 | 0.06 |
| tapentadol (J) | -0.03 | 0.21 | 0.03 | 0.05 | 0.34 | -0.02 | 0.02 | 0.43 | 0.49 | 0.17 | 0.07 |

## Discussion

There are several novel findings from this investigation on the dynamic changes and inter-state variance in the distribution of prescription opioids in the US. This study determined that distribution of prescription opioids for pain has undergone a pronounced reduction over the past decade, which is congruent with and extends upon reports from earlier periods [2-6, 11]. This report identified a consistent decline in opioid distribution since 2011 [3]. There are a myriad of reasons that may explain this decline, including increased awareness of the addictive nature of opioids [22], continued escalation in opioid-related overdose mortalities [1], and the increased government funding and resources resulting from the classification of the opioid crisis as a public health emergency [23]. Evidence has been more subtle or contradictory for any measurable impact of Prescription Drug Monitoring Programs [24, 25] or state opioid prescribing laws [12, 15, 26].

Oxycodone distribution was the most pronounced relative to the other nine opioids and also demonstrated the largest decline in opioid distribution relative to its peak year. This appreciable decline could be due to factors such as pharmaceutical market changes as exemplified in the 2010 introduction of abuse-deterrent OxyContin. Two years after the 2010 formulation change, the dispensing rate for extended-release oxycodone decreased by 39% [27].

Although there has been much prior attention to opioid distribution [2-6, 8-14], the changes in the production quotas have received limited quantitative attention. The total opioid production quota from 2010 to 2019 demonstrated a pronounced increase from 2013 until 2016. There was a forty-percent decline after 2016. The DEA is required to set these aggregate production quotas (APQ), one of three types of quotas set by the Administration, each year at levels that provide for the “estimated medical, scientific, research and industrial needs of the United States” and related purposes [28]. As the Department of Justice’s Office of the Inspector General describes in a 2019 report, the DEA consistently failed to ensure that the quotas for many opioids were consistent with actual medical need for the drugs, and in 2017 administratively reduced the APQ for most controlled substances by 25% [29]. The DEA modified its regulations in August 2018 to change the factors for the DEA to consider in making determinations about the quota, including “the extent of any diversion of the controlled substance in the class” as well as relevant information from FDA, CDC, and the states [30].

The total production quotas (Figure 1A) were about three-fold higher than opioid distribution (Figure 1B) throughout the study period, suggesting that the quotas had little to no impact on distribution of those medications and were not based on a systematic evaluation of medical need. One caveat is that distribution only included methadone when used for pain while the production quota included methadone that was subsequently employed for both pain and narcotic treatment programs. By MME, methadone is by far the predominant prescription opioid in the US [3]. However, quotas for other opioid drugs such as hydrocodone, oxycodone, and oxymorphone also increased from 2013 to 2016, even though distribution of those drugs decreased every year during that period, demonstrating a large disconnect between the production quotas and actual distribution.

Although there have been overall reductions in opioid distribution, each opioid experienced varying degrees of distribution in a given year. The high-potency opioid fentanyl demonstrating the lowest variation (2011 percentile ratio = 1.77, 2019 ratio = 2.12) which extends upon prior reports [10,11]. Methadone rose to have the highest degree of variation in 2019 despite being 3rd highest in 2011. The observed variation can be due to the lack of federal quantity limits on Schedule II drug prescriptions, which allows individual states and health insurance carriers to impose their own limitations regarding prescribing and dispensing Schedule II drugs. These restrictions on drug dispensation vary greatly on bases such as limitations on drug days’ supply, dosage unit, and whether the restrictions apply to only the first prescription [31,32]. Further, formulation changes, such as that seen with oxycodone [27], could play a role in the observed differences in degree of variation in distribution for a particular opioid in a particular year. However, the federal reclassification of hydrocodone from a Schedule III to a Schedule II drug in 2014 [33] did not appreciably impact the variation (2011 percentile ratio = 6.11, 2019 ratio = 7.99).

Similarly, opioid distribution was not homogeneous. Total per-capita opioid distribution from 2011 was significantly elevated in Tennessee, Nevada, Delaware, and Florida relative to the national mean. When analyzing the total per capita opioid distribution in 2019, only two states, Alabama and Delaware, were significantly higher than the national average. Further, when analyzing the data nationally, Florida [8] had the highest per capita opioid distribution decline while Texas [34] had the lowest decline. We can speculate that the general allowance of state discretion on what policies to implement and how to implement/enforce them may play a role in these observed differences in progress towards decreasing opioid distribution.

There were several positive correlations between opioid distribution per person per state. Interestingly, both 2011 and 2019 showed multiple associations with oxycodone. This finding suggests that there may be a link between the tendency of being prescribed oxycodone alongside another opioid medication. Strong correlations were also found for codeine and hydrocodone, codeine and hydromorphone, hydrocodone and methadone, and methadone and oxycodone. It is very likely that medical specialty preferences exist for the choice of opioid medication based on knowledge of opioid pharmacology, comfort level, and experience. Additional studies should explore the influence on treatment decisions at a patient level to further explore these opioid associations.

While we identified changes in Schedule II opioid production quotas and distribution between 2011 and 2019 and greatly extended upon prior investigations [2, 3, 8-13, 23, 34] to explore the substantial state and regional variation in opioid distribution [36], this study was not without caveats. We reported a decline in opioid distribution beginning in 2011 based on the distribution rates of the opioids as mentioned earlier. However, another commonly prescribed opioid, buprenorphine [13] was not included in this analysis and may have influenced to a small degree the peak year of opioid distribution. In addition, ARCOS data does not account for Schedule IV or V opioids that could influence the total opioid distribution across the US [37]. For example, tramadol was moved from a non-controlled medication to Schedule IV in 2014 [38]. The MMEs per capita is for opioids primarily employed for pain would be even higher with the inclusion of additional non-Schedule II opioids. Moreover, the effects of racial, gender and socioeconomic disparities were not examined here but have been explored by others [39-43]. Stratifying data based on these demographics could identify more specific changes and further characterize continued disparities in opioid distribution.

Overall, this study found dynamic changes in prescription opioid production quotas and sizable but gradual reductions in distribution across the US from 2010 to 2019. The production quotas seem to have little to no correspondence with medical need for opioid medications as expressed in the actual amount of opioids distributed. Further, the non-homogenous nature of opioid distribution and its effects on the population pose a barrier to continued efforts to one-day overcoming the US opioid epidemic. Further investigations of state and county level differences may inform the policies of opioid stewardship programs.

## Data Availability

All data produced are available online at:
https://www.deadiversion.usdoj.gov/arcos/retail_drug_summary/

## Acknowledgments

We would like to thank Geisinger Commonwealth School of Medicine for supporting this research and the Biomedical Research Club for technical assistance. Lori Heagle, MFA provided technical assistance preparing figures. We thank the Drug Enforcement Administration for making the Automated Reports and Consolidated Orders System dataset publicly available.

---

**Supplemental Figure 1.**
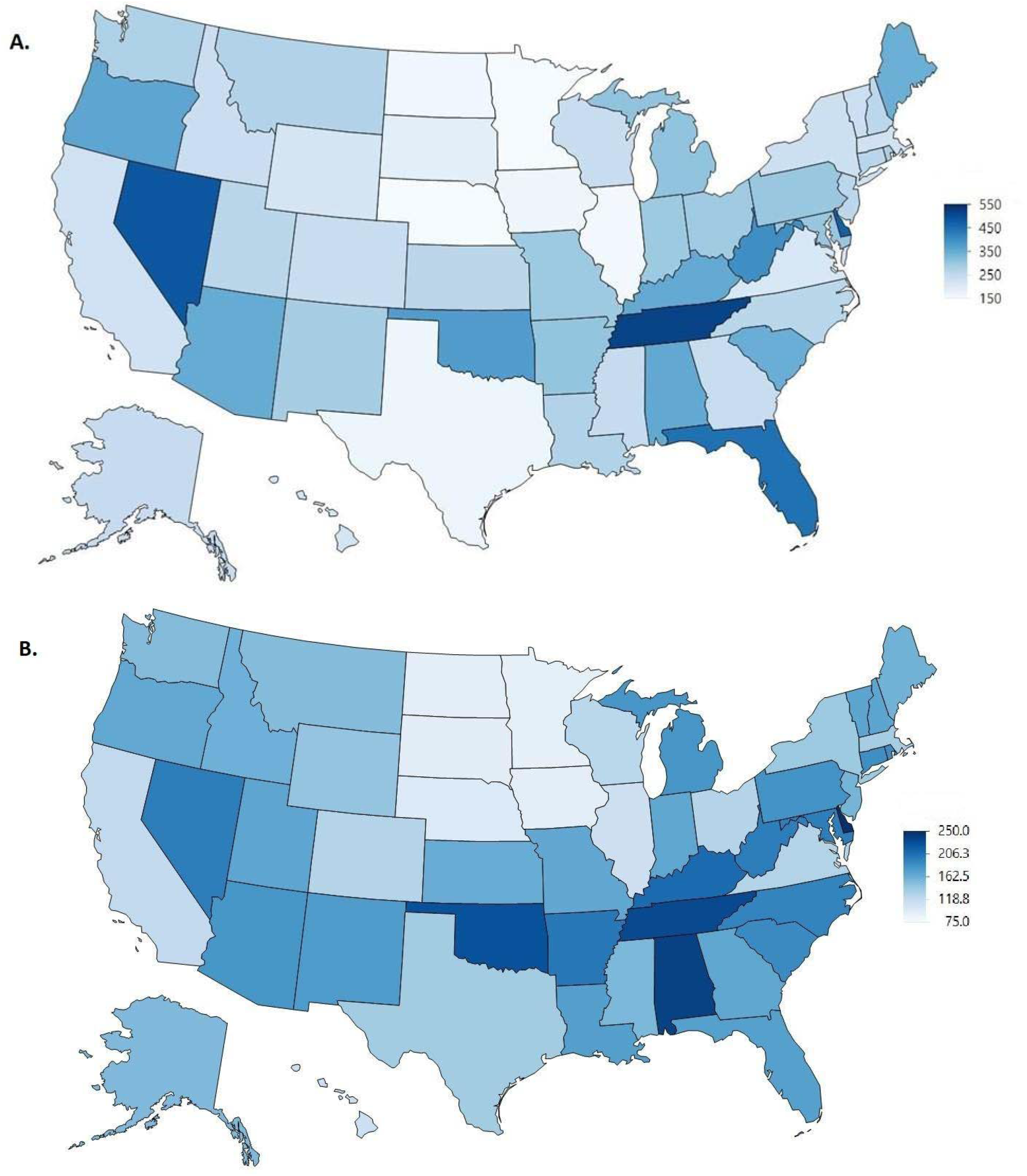
Heat maps of the total opioid distribution in morphine mg equivalents (MME) per person across the United States in 2011 (A) and 2019 (B).

**Supplemental Table 1.** US Drug Enforcement Administration production data quotas.

| Year | Source |
| --- | --- |
| 2010 | https://www.deadiversion.usdoj.gov/fed_regs/quotas/2010/fr0914.htm |
| 2011 | https://www.deadiversion.usdoj.gov/fed_regs/quotas/2011/fr1209.htm |
| 2012 | https://www.deadiversion.usdoj.gov/fed_regs/quotas/2012/fr0910.htm |
| 2013 | https://www.deadiversion.usdoj.gov/fed_regs/quotas/2013/fr0807.htm |
| 2014 | https://www.deadiversion.usdoj.gov/fed_regs/quotas/2014/fr0825.htm |
| 2015 | https://www.deadiversion.usdoj.gov/fed_regs/quotas/2015/fr0916.htm |
| 2016 | https://www.deadiversion.usdoj.gov/fed_regs/quotas/2016/fr1025.htm |
| 2017 | https://www.deadiversion.usdoj.gov/fed_regs/quotas/2017/fr1103.htm |
| 2018 | https://www.deadiversion.usdoj.gov/fed_regs/quotas/2018/fr1210.htm |
| 2019 | https://www.deadiversion.usdoj.gov/fed_regs/quotas/2019/fr1202.htm |

**Supplemental Table 2.** Distribution of Schedule II opioids as reported by the Drug Enforcement Administration’s Automated Reports and Consolidated Orders System (ARCOS), corrected for population according to the annual American Community Survey, in 2011 (top) and 2019 (bottom). The abbreviations of the highest and lowest states are in superscript.

| **2011** | <u>70<sup>th</sup></u><br><u>Percentile</u> | <u>30<sup>th</sup></u><br><u>Percentile</u> | <u>70<sup>th</sup> : 30<sup>th</sup></u><br><u>Ratio</u> | <u>Maximum</u><br><u>State</u> | <u>Minimum</u><br><u>State</u> | <u>Max : Min</u><br><u>Ratio</u> |
| --- | --- | --- | --- | --- | --- | --- |
| codeine | 28.94 | 22.70 | 1.27 | 57.70<sup>MI</sup> | 14.28<sup>NY</sup> | 4.04 |
| fentanyl | 0.92 | 0.70 | 1.32 | 1.17<sup>SD</sup> | 0.47<sup>HI</sup> | 2.50 |
| hydrocodone | 78.77 | 43.80 | 1.80 | 163.93<sup>TN</sup> | 17.78<sup>NJ</sup> | 9.22 |
| hydromorphone | 2.72 | 2.01 | 1.35 | 4.84<sup>DE</sup> | 1.05<sup>HI</sup> | 4.61 |
| meperidine | 4.03 | 2.23 | 1.81 | 13.89<sup>AR</sup> | 0.71<sup>MN</sup> | 19.56 |
| methadone | 29.70 | 17.16 | 1.73 | 64.57<sup>ME</sup> | 5.79<sup>SD</sup> | 11.15 |
| morphine | 44.63 | 31.99 | 1.40 | 81.88<sup>OR</sup> | 19.77<sup>TX</sup> | 4.14 |
| oxycodone | 122.84 | 84.30 | 1.46 | 304.57<sup>DE</sup> | 22.44<sup>IL</sup> | 13.57 |
| oxymorphone | 3.31 | 1.74 | 1.91 | 10.42<sup>TN</sup> | 0.38<sup>MN</sup> | 27.42 |
| tapentadol | 8.59 | 4.58 | 1.87 | 19.60<sup>AL</sup> | 1.40<sup>MN</sup> | 14.00 |

| **2019** | <u>70<sup>th</sup></u><br><u>Percentile</u> | <u>30<sup>th</sup></u><br><u>Percentile</u> | <u>70<sup>th</sup> : 30<sup>th</sup></u><br><u>Ratio</u> | <u>Maximum</u><br><u>State</u> | <u>Minimum</u><br><u>State</u> | <u>Max : Min</u><br><u>Ratio</u> |
| --- | --- | --- | --- | --- | --- | --- |
| codeine | 14.48 | 10.71 | 1.35 | 57.63<sup>TX</sup> | 5.87<sup>CO</sup> | 9.82 |
| fentanyl | 0.36 | 0.28 | 1.29 | 0.55<sup>MS</sup> | 0.18<sup>HI</sup> | 3.05 |
| hydrocodone | 38.49 | 20.38 | 1.89 | 78.39<sup>AL</sup> | 5.84<sup>NJ</sup> | 13.42 |
| hydromorphone | 1.64 | 1.24 | 1.32 | 3.41<sup>MS</sup> | 0.70<sup>HI</sup> | 4.87 |
| meperidine | 0.66 | 0.33 | 1.99 | 2.48<sup>AK</sup> | 0.11<sup>RI</sup> | 22.17 |
| methadone | 28.56 | 15.68 | 1.82 | 86.88<sup>RI</sup> | 2.87<sup>WY</sup> | 30.22 |
| morphine | 22.94 | 16.33 | 1.40 | 34.33<sup>TN</sup> | 10.85<sup>TX</sup> | 3.16 |
| oxycodone | 68.83 | 53.59 | 1.28 | 109.65<sup>DE</sup> | 17.89<sup>TX</sup> | 6.13 |
| oxymorphone | 0.69 | 0.32 | 2.16 | 2.71<sup>TN</sup> | 0.07<sup>MN</sup> | 38.92 |
| tapentadol | 6.64 | 3.94 | 1.68 | 15.28<sup>NC</sup> | 1.24<sup>MN</sup> | 12.31 |

